# Targeted Proteomics of Right Heart Adaptation to Pulmonary Arterial Hypertension

**DOI:** 10.1101/2020.06.19.20136028

**Authors:** Myriam Amsallem, Andrew J. Sweatt, Jennifer Arthur Ataam, Julien Guihaire, Florence Lecerf, Mélanie Lambert, Maria Rosa Ghigna, Md Khadem Ali, Yuqiang Mao, Elie Fadel, Marlene Rabinovitch, Vinicio de Jesus Perez, Edda Spiekerkoetter, Olaf Mercier, Francois Haddad, Roham T. Zamanian

## Abstract

**Rationale:** No prior proteomic screening study has centered on the right ventricle (RV) in pulmonary arterial hypertension (PAH).

**Objectives:** To identify the circulating proteomic profile associated with right heart maladaptive phenotype (RHMP) in PAH.

**Methods:** Plasma proteomic profiling was performed using multiplex immunoassay in 121 PAH patients (discovery cohort from 2008-2011) and 76 (validation cohort from 2012-2014). The association between proteomic markers and RHMP (defined by the Mayo right heart score [combining RV strain, NYHA and NT-proBNP] and Stanford score [RV end-systolic remodeling index, NYHA and NT-proBNP]) was assessed by partial least squares regression. Expression levels of biomarkers were measured in RV samples from PAH patients undergoing transplant and controls, and mice subjected to pulmonary artery banding (PAB).

**Measurements and Main Results:** High levels of hepatic growth factor (HGF), stem cell growth factor beta, nerve growth factor and stromal derived factor-1 were significantly associated with worse Mayo and Stanford scores but not with pulmonary vascular resistance or pressure in both discovery and validation cohorts (this latter had more severe disease features: lower cardiac index and higher NT-proBNP). In both cohorts, HGF added incremental value to the REVEAL score in the prediction of death, transplant, or hospitalization at 3 years. RV expression levels of HGF and its receptor c-Met were higher in end-stage PAH patients than controls, and in PAB mice than shams.

**Conclusion:** High plasma HGF levels are associated with a RHMP and predictive of 3-year clinical worsening. Both HGF and c-Met RV expression levels are increased in PAH.

**At a Glance Commentary:** *Scientific Knowledge on the Subject:* Right heart maladaptation is the main cause of death in patients with pulmonary arterial hypertension (PAH). Recent non-invasive imaging studies have improved right heart adaptive phenotyping in PAH, identifying right ventricular (RV) free-wall longitudinal strain and the RV end-systolic remodeling index as markers of right heart maladaptive phenotypes (RHMP). Emerging evidence suggests a link between inflammation and RV failure in PAH patients and experimental disease models, yet no prior proteomic screening study has centered on the right heart.

*What This Study Adds to the Field:* This targeted proteomics screening study identifies 4 plasma biomarkers of RHMP in two clinical cohorts of patients with PAH. Among them, hepatic growth factor is shown to be prognostic and incremental to the REVEAL risk score for prediction of 3-year death, lung transplant and readmission. HGF and its receptor c-Met are overexpressed in RV tissue samples from PAH patients undergoing transplant as compared to controls, and mice subjected to pulmonary artery banding (PAB), warranting further exploration as a potential right ventricular-specific therapeutic target.

*Online data supplements:* This article has an online data supplement, which is accessible from this issue’s table of content online at www.atsjournals.org.

## INTRODUCTION

Pulmonary arterial hypertension (PAH) is characterized by progressive obliterative vascular remodeling, which leads to increased right heart afterload. The right ventricle (RV) initially adapts by increasing its wall thickness (adaptive phenotype), but PAH progression often results in RV enlargement and failure (maladaptive phenotype) with subsequent death or lung transplantation (1). The pathophysiology underlying this transition from an adapted to maladapted RV is not well elucidated, in part because very few PAH research efforts have focused on establishing markers of the right heart maladaptive phenotype (RHMP). Previous proteomic studies of PAH have uncovered blood profiles associated with pulmonary vascular disease severity and outcomes, including our own work, which utilized unsupervised machine learning to identify PAH phenotypes with distinct inflammatory profiles that stratify clinical risk (2, 3). Emerging evidence suggests a link between inflammation and RV failure in PAH patients and experimental disease models (4, 5), yet no prior proteomic screening study has centered on the right heart.

Recent imaging studies have substantially improved right heart adaptive phenotyping in PAH, identifying RV free-wall longitudinal strain (RVLS) and end-systolic dimensions (such as the remodeling index RVESRI) as strong echocardiographic markers of RV adaptation (6, 7). These markers have each been integrated with New York Heart Association (NYHA) functional class and N-terminal pro B-type natriuretic peptide (NT-proBNP) to define RHMP scores – a Mayo Clinic model (based on RVLS, NYHA class and NT-proBNP) (6) and a Stanford model (RVESRI, NYHA class and NT-proBNP) (7). The prognostic value of these simple RV-centered models is equivalent to the REVEAL score (7), a widely adopted PAH risk stratification tool, which requires more extensive clinical data and invasive hemodynamic measures.

Building upon these right heart scores, our primary objective was to identify novel circulating proteomic biomarkers of RHMP in PAH. We hypothesized that PAH patients may express a specific blood proteomic profile in association with RHMP, independent of pulmonary disease severity. We also aimed to examine the relationship of RHMP proteomic markers to survival. Finally, we sought to assess tissue-level expression of the RHMP markers in RV samples from end-stage PAH patients and an experimental mouse model of RV pressure overload, to test the hypothesis that increased circulating levels reflect RV overexpression, and to also validate the PAB model for future mechanistic studies.

## METHODS

### Study population

This prospective study included group 1 PAH patients enrolled into the Stanford University Pulmonary Hypertension Biobank (Stanford, CA) between 2008 and 2014, who had plasma collected for proteomic profiling within two weeks of a routine echocardiogram (**Figure 1, Supplementary Methods**). PAH diagnosis required mean pulmonary arterial pressure (MPAP) ≥25 mmHg, pulmonary arterial wedge pressure ≤15 mmHg, and pulmonary vascular resistance (PVR) >240 dyn.s.cm^-5^. Exclusion criteria were: chronic infection (n=7), primary immunodeficiency (n=1), recent acute illness other than heart failure (±1 month) (n=8), congenital systemic-to-pulmonary shunt (n=10), active malignancy (n=4), missing echocardiogram within 2 weeks (n=68), or suboptimal echocardiographic data (n=6). Study baseline was the date of blood collection. Patients were divided into a discovery (n=121, 2008-2011) and validation cohort (n=76, 2012-2014). Plasma was also obtained from healthy controls (n=88) who underwent rigorous screening to establish health. Stanford Institutional Review Board approved the study (IRB #14083 and #20942); all subjects provided informed consent.

**Figure 1.**
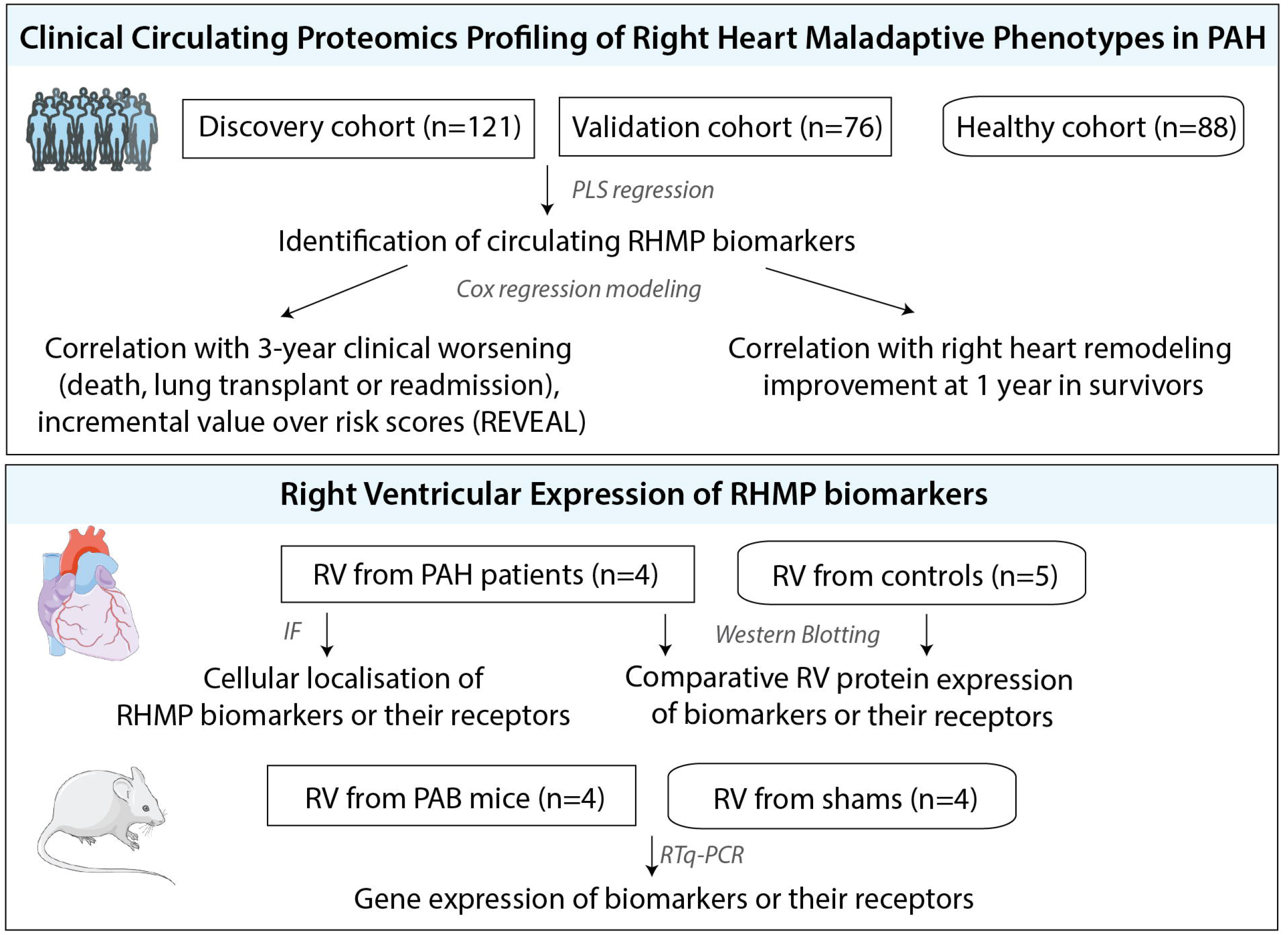
Study design. PAB: pulmonary arterial banding; PAH: pulmonary artery hypertension; RHMP: right heart maladaptive phenotype.

### Blood proteomic profiling

Fasting blood samples were collected from an antecubital vein, processed to plasma, and stored according to protocols (**Supplementary Methods**). A targeted proteomic panel of 48 cytokines, chemokines and growth factors (**Table 1** and **Table E1**) was measured for each subject using Bio-Plex multiplex immunoassay (BioRad Inc., Hercules, CA), a bead-based flow cytometric platform built on Luminex® xMAP (tm) technology (Luminex Corp., Austin, TX), according to manufacturer instructions (**Supplementary Methods**). Each sample was measured in duplicate. A Luminex 200 plate reader quantified median fluorescence intensity (MFI) for each protein. MFI was used for all analyses rather than standard curve-derived absolute concentrations, as MFI does not require detection limit censoring and has better downstream statistical power (8). The inter-assay coefficient of variation was <15% for all measured analytes, based on internal controls included across runs.

**Table 1.**
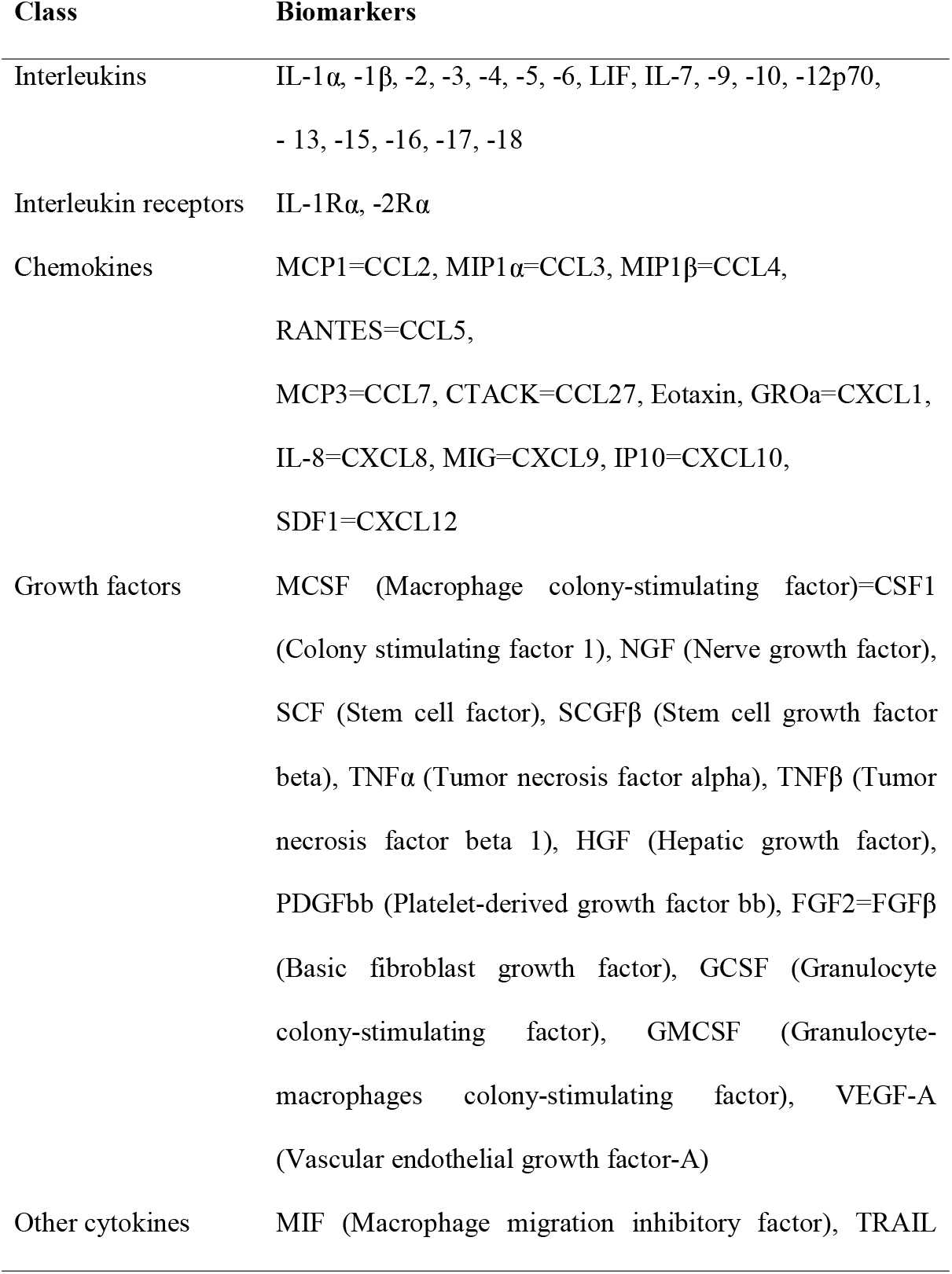
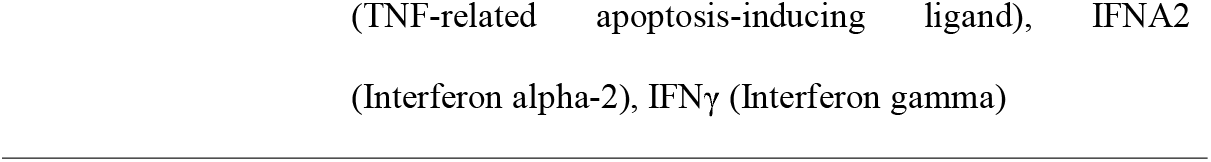
Circulating proteomic biomarkers assessed in the study using flow cytometry multiplex arrays.

| **Class** | **Biomarkers** |
| --- | --- |
| Interleukins | IL-1α, -1β, -2, -3, -4, -5, -6, LIF, IL-7, -9, -10, -12p70,<br>- 13, -15, -16, -17, -18 |
| Interleukin receptors | IL-1Rα, -2Rα |
| Chemokines | MCP1=CCL2, MIP1α=CCL3, MIP1β=CCL4,<br>RANTES=CCL5,<br>MCP3=CCL7, CTACK=CCL27, Eotaxin, GROα=CXCL1,<br>IL-8=CXCL8, MIG=CXCL9, IP10=CXCL10,<br>SDF1=CXCL12 |
| Growth factors | MCSF (Macrophage colony-stimulating factor)=CSF1<br>(Colony stimulating factor 1), NGF (Nerve growth factor),<br>SCF (Stem cell factor), SCGFβ (Stem cell growth factor<br>beta), TNFα (Tumor necrosis factor alpha), TNFβ (Tumor<br>necrosis factor beta 1), HGF (Hepatic growth factor),<br>PDGFbb (Platelet-derived growth factor bb), FGF2=FGFβ<br>(Basic fibroblast growth factor), GCSF (Granulocyte<br>colony-stimulating factor), GMCSF (Granulocyte-<br>macrophages colony-stimulating factor), VEGF-A<br>(Vascular endothelial growth factor-A) |
| Other cytokines | MIF (Macrophage migration inhibitory factor), TRAIL |

### Clinical data collection

Transthoracic echocardiogram studies were acquired using Philips IE 33 ultrasound systems (Philips, Amsterdam, Netherlands). RV dimensions and functional metrics were measured on RV-focused apical 4-chamber views, averaged over three cycles, and included RVESRI (defined by the lateral wall to septal height ratio) and RVLS as previously published (**Supplementary Methods**) (7). These measurements were used to calculate two right heart adaptive phenotyping scores the Mayo Clinic score integrating RVLS, NYHA class and NT-proBNP, and the Stanford score integrating RVESRI, NYHA class and NT-proBNP (**Table E2**). Follow-up echocardiograms available in survivors at 1 year were interpreted using the same methodology. Right heart remodeling with therapy was defined by changes in RVESRI from baseline (improvement if relative delta <10%, worsening if >10% and stable otherwise).

NYHA class, six-minute walk distance, diffusing capacity of the lung for carbon monoxide, MDRD estimated glomerular filtration rate and NT-proBNP levels were collected if available within 1 month of baseline. Hemodynamic data (right atrial pressure, MPAP, PAWP, PVR, and cardiac index) were obtained when right heart catheterization was performed within 3 months of inclusion. Data were used to calculate the REVEAL risk score (**Table E3**) (9).

Patients were prospectively followed to evaluate the primary endpoint of clinical worsening (i.e. death, lung transplantation, or hospitalization for acute right heart failure), and the secondary end point (death or lung transplant).

### Pathology analysis of human RV samples

RV frozen and paraffin-embedded biopsies of explanted hearts were obtained from 4 PAH patients (**Table E4**) who underwent heart-lung transplant at Marie Lannelongue Hospital (IRB #18.06.06, Le Plessis-Robinson, France). For comparison, we acquired RV frozen biopsies from 5 control heart donors (without cardiovascular disease or pulmonary hypertension) whose organs were not transplanted (paraffin-embedded biopsies were not available); Explanted hearts were procured from the INSERM URMS 1148 biobank (BRIF BB-0033-00029, BBMRI-EU/infrastructure), approved by the French National Ethics Committee (N°: PFS17-002) and declared at the French Ministry of Research (DC-2018-3141).

Upon identifying RHMP circulating biomarkers in the clinical cohorts, we (i) compared tissue-level expression of these proteins and their receptors in PAH and controls by incubating frozen RV samples with primary specific antibodies (Abcam, Cambridge, UK) and performing western blots, and (ii) localized the RHMP biomarkers in RV tissue from PAH patients with immunofluorescence staining and confocal microscopy (Zeiss LSM 800 microscope with ZEN software; Carl Zeiss Microscopy, White Plains, NY) (**Supplementary Methods**).

### Pulmonary Artery Banding (PAB) mouse model RV analysis

Chronic RV pressure overload was induced in 10-14 week-old male C57BL/6 mice (**Supplementary Methods**) (10). Anesthetized animals underwent thoracotomy, 6-0 silk sutures were used to band the main pulmonary artery (PA), and one-week post-surgery a peak pressure gradient >15 mmHg was confirmed across the band by echocardiography (GE Vivid 7). A control group of age-matched littermates underwent sham surgery (PA isolation without suture placement). MPAP, heart rate, and pulmonary valve velocity time integral were measured by echocardiography after 5 weeks to document reduced cardiac output and RV samples were collected and snap frozen. After isolating RNA from homogenized RV samples, we applied quantitative reverse transcriptase-PCR (Applied Biosystems, Foster City, CA) to measure expression levels of the mRNA transcripts corresponding to RHMP proteomic markers of interest.

### Statistical analysis

Baseline characteristics were compared using Student’s t-test or Mann-Whitney-Wilcoxon test for continuous data, and Chi-square test for categorical data. Proteomic data preprocessing involved background fluorescence subtraction, plate/batch adjustment (empirical Bayes methodology) (11), robust quantile normalization, and duplicate averaging, and adjustment for age, sex, and body mass index. To identify RHMP markers, partial least squares (PLS) regression models were fit using the ‘SIMPLS’ algorithm (**Supplementary Methods**). RHMP markers were selected by assessing (i) variable importance for projections (VIP) scores and (ii) regression beta coefficients (using t-tests of coefficients based on variance estimates from jackknife resampling). PLS models were fit to associate proteins with Mayo and Stanford scores, individual components of each score, and pulmonary hemodynamics. Levels of the identified RHMP biomarkers were examined among low, intermediate, and high-risk RV phenotypes (based on Mayo and Stanford scores) to compare each risk group to controls (Kruskal-Wallis and post-hoc Dunn tests) and confirm a trend across ordered groups (Cuzick test). To relate biomarkers to outcomes, Kaplan-Meier estimates of transplant- and hospitalization-free survival were compared across quartiles by log-rank statistics, and Cox proportional hazards regression models were fit. Scaled Schoenfeld residuals were plotted against time for each variable, to ensure the proportional hazards assumption was met. Hazard ratios were normalized to the standard deviation of predictor variables. To determine whether each biomarker added incremental value to established risk scores for prediction of 3-year outcome, χ2 values were compared. Cox regression model was used to determine whether biomarkers or right heart metrics were predictive of RVESRI improvement at 1 year. Patients included in 2014 with <3 years follow-up (n=4) were right censored. Statistical analyses were performed using SPSS® v.19 (SPSS Inc, Chicago, IL) and R v.3.5.1 (R Core Team, Vienna, Austria) software programs.

## RESULTS

### Proteomic profiling of RHMP in patients with PAH

**Table 2** summarized baseline characteristics of the discovery cohort. High levels of hepatic growth factor (HGF), stem cell growth factor (SCGFβ or CLEC11A), nerve growth factor (NGF) and stromal cell-derived factor 1 (SDF1 or CXCL12) were associated with higher-risk Mayo and Stanford scores, RV dysfunction (RVLS), RV adverse remodelling (RVESRI), higher NT-proBNP and worse NYHA functional class, but not with pulmonary disease severity (**Figure 2A, Table E5** and **Figure E1)**.

**Table 2.** Comparative baseline characteristics of the discovery and validation cohorts.

| **Variables** | **Discovery cohort**<br>**n=121** | **Validation cohort**<br>**n=76** | **p value** |
| --- | --- | --- | --- |
| Age (years) | 50 [39 – 59] | 52 [43 – 61] | 0.20 |
| Female sex | 90 (74.4) | 66 (86.8) | **0.04** |
| Body mass index (kg/m<sup>2</sup>) | 26.9 [22.5 – 31.9] | 27.4 [24.7 – 33.5] | 0.12 |
| PAH etiology |  |  | 0.40 |
| Connective tissue disease | 40 (33.1) | 31 (40.8) |  |
| Idiopathic | 34 (28.1) | 14 (18.4) |  |
| Drugs and toxins | 21 (17.4) | 18 (23.7) |  |
| Congenital heart disease | 13 (10.7) | 7 (9.2) |  |
| Portal pulmonary hypertension | 11 (9.1) | 5 (6.6) |  |
| Heritable PAH | 2 (1.7) | 1 (1.3) |  |
| NYHA functional class |  |  | 0.26 |
| Class I | 8 (6.6) | 5 (6.6) |  |
| Class II | 48 (39.7) | 20 (26.3) |  |
| Class III | 50 (41.3) | 41 (53.9) |  |
| Class IV | 15 (12.4) | 10 (13.2) |  |
| Six minute walk distance (m) | 427 [342 – 537] | 392 [299 – 476] | **0.03** |
| NT-proBNP (pg/mL) | 235 [67 – 977] | 536 [150 – 1441] | **0.04** |
| DLCO (% predicted) | 73 [60 – 86] | 69 [53 – 92] | 0.91 |
| GFR (mL/min/1.73m<sup>2</sup>) | 68 [53 – 90] | 66 [54 – 79] | 0.21 |
| **Hemodynamics** |  |  |  |
| Right atrial pressure (mmHg) | 7 [5 – 11] | 7 [5 – 12] | 0.49 |
| Mean pulmonary arterial pressure (mmHg) | 50 [40 – 59] | 49 [40 – 58] | 0.78 |
| Pulmonary arterial wedge pressure (mmHg) | 10 [8 – 13] | 10 [7 – 14] | 0.72 |
| Pulmonary vascular resistance (WU) | 9.9 [6.1 – 14.2] | 10.3 [6.8 – 15.7] | 0.56 |
| Cardiac Index (L/min/m<sup>2</sup>) | 2.2 [1.8 – 2.5] | 2.0 [1.6 – 2.3] | **0.03** |
| **Echocardiographic metrics** |  |  |  |
| RVESAI (cm<sup>2</sup>/m<sup>2</sup>) | 12.7 [10.5 – 16.4] | 12.9 [10.2 – 16.2] | 0.81 |
| RVESRI | 1.4 [1.3 – 1.6] | 1.5 [1.3 – 1.6] | 0.43 |
| RVLS (absolute value, %) | 16.9 [13.9 – 20.0] | 16.6 [13.2 – 21.1] | 0.93 |
| RVFAC (%) | 27.1 [22.7 – 30.7] | 26.6 [23.1 – 32.8] | 0.91 |
| TAPSE (cm) | 1.8 [1.4 – 2.1] | 1.7 [1.3 – 2.0] | 0.20 |
| **PAH Therapy** |  |  |  |
| Therapy extent |  |  | **<0.01** |
| Treatment naïve | 35 (28.9) | 38 (50.0) |  |
| Monotherapy | 34 (28.1) | 24 (31.6) |  |
| Dual therapy | 36 (29.8) | 12 (15.8) |  |
| Triple therapy | 16 (13.2) | 2 (2.6) |  |
| Phosphodiesterase-5 inhibitors | 62 (51.2) | 28 (36.8) | **0.04** |
| Endothelin receptor antagonists | 39 (32.2) | 12 (15.8) | **0.01** |
| Prostanoid therapy | 53 (43.8) | 14 (18.4) | **<0.01** |
| **REVEAL Risk Score** |  |  | **0.48** |
| Low | 74 (61.2) | 46 (60.5) |  |
| Average/Moderate High Risk | 17 (14.0) | 15 (19.7) |  |
| High/Very High | 30 (24.8) | 15 (19.7) |  |
| **Mayo Right Heart Score** |  |  | **0.85** |
| Low score (0-5) | 50 (41.3) | 33 (43.3) |  |
| Intermediate score (6-8) | 40 (33.1) | 28 (36.8) |  |
| High score (>9) | 24 (19.8) | 15 (19.7) |  |
| **Stanford Right Heart Score** |  |  | **0.72** |
| Low score (0-2) | 49 (40.5) | 30 (39.5) |  |
| Intermediate score (3) | 46 (38.0) | 26 (34.2) |  |
| High score (>4) | 26 (21.5) | 20 (26.3) |  |

Values are expressed as mean ± SD or number (percentage), or median and interquartile range (IQ). Significant p values ( $<0.05$ ) are presented as bold. RVSP was estimable from the TR signal in 194 (85% patients). NYHA: New York Heart Association; PAH: pulmonary arterial hypertension; RV: right ventricular; RVESAI: RV end-diastolic area indexed on body surface area; RVFAC: fractional area change; RVLS: free-wall longitudinal strain; RVSP: right ventricular systolic pressure; TAPSE: tricuspid annular plane systolic excursion.

**Figure 2.**
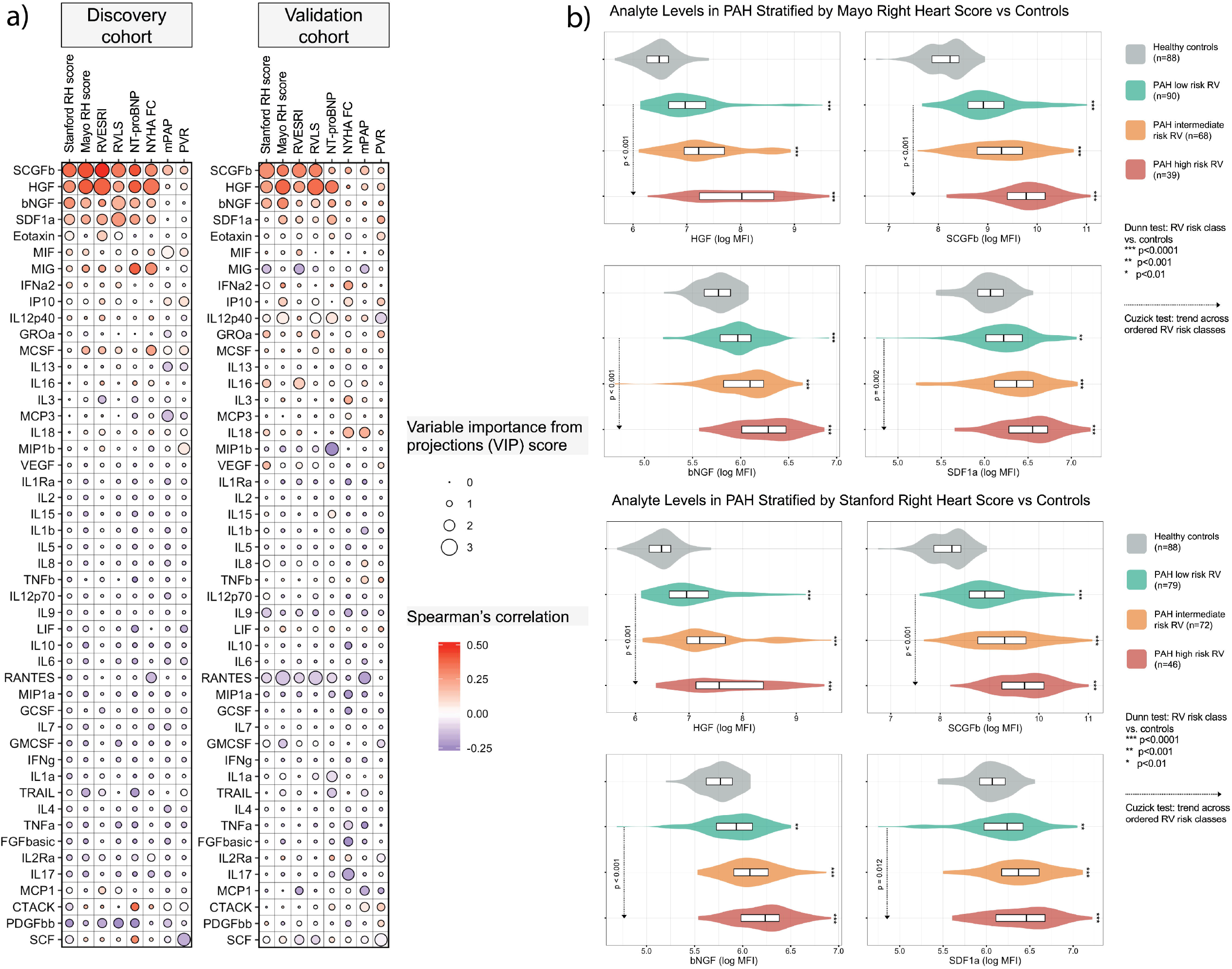
(a) Variable Importance in Projection (VIP) scores and correlations for each cytokine with respect to right heart scores and metrics in both cohorts and (b) Levels of the 4 markers according to levels of right heart maladaptive phenotype in patients with pulmonary arterial hypertension (PAH) and controls. **(a)** Using partial least square regression, we assessed the correlations between plasma proteomic markers (using multiplex immunoassay) and markers of right heart maladaptive phenotype (Stanford right heart RH score based on RVESRI, NYHA class and NT-proBNP levels; Mayo RH score based on RVLS, NYHA class and NT-proBNP levels; RVESRI; RVLS; NT-proBNP; NYHA FC; mPAP and PVR) in 121 PAH patients (discovery cohort from 2008-2011) and 76 (validation cohort from 2012-2014). Positive correlations are shown in red, negative in blue. **(b)** Analyte levels of the 4 strongest biomarkers associated with right heart maladaptive phenotype according to tertiles of the Stanford RH score (upper panel) or Mayo RH score (lower panel) in patients with PAH and in the 88 healthy controls. MPAP: mean pulmonary arterial hypertension; NYHA FC: New York Heart Association functional class; PVR: pulmonary vascular resistance; RH: right heart; RVLS: right ventricular free-wall longitudinal strain is presented in absolute value (lowest values indicate worst right ventricular dysfunction); RVESRI: right ventricular end-systolic remodeling index. For cytokines abbreviations, please see **Table 1** and supplementary **Table S1**.

The validation cohort had a greater proportion of females, more severe pulmonary disease features, fewer patients on background therapy and a higher frequency of incident cases than the discovery cohort (**Table 2**). REVEAL, Mayo and Stanford risk scores distributions were similar in the two cohorts. High levels of HGF, SCGFβ, NGF, and to a lesser extent SDF1, were associated with higher-risk Mayo and Stanford scores, RV dysfunction, higher NT-proBNP and worse NYHA functional class, but not with pulmonary disease severity (**Figure 2A**). No measured cytokine was found to have a positive association with favorable right heart metrics across both cohorts.

**Figure 2B-C** compared the 4 candidate RHMP markers levels in both cohorts combined among low, intermediate, and high-risk RV patient subgroups, and the 88 healthy controls (median age 58 [44-64] years, 52.3% were female).

### Prognostic value of HGF

During a median follow-up of 3.14 [2.16; 5.44] years, the primary end point was reached in 76 patients (58 were hospitalized, 10 transplanted and overall 41 died). Event-free survival rates (± standard error) for the primary end point were 91 ±2% at 1 year, 78 ±3% at 3 years, and 62 ±34% at 5 years (**Figure 3A**). The secondary end point of death or transplant was reached in 49 patients.

**Figure 3.**
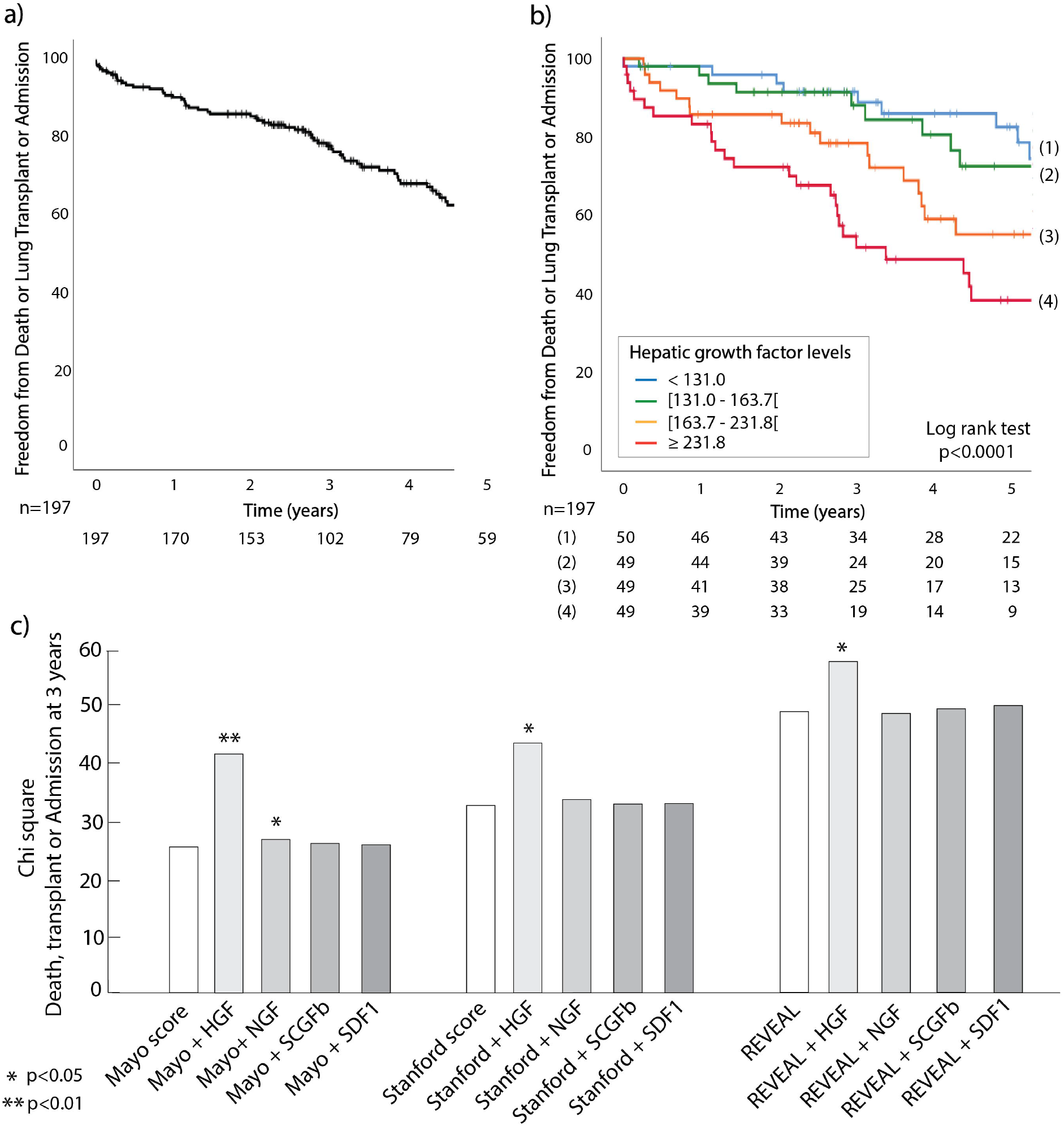
Prognostic value of hepatic growth factor (HGF) and its incremental value to risk scores for outcome prediction. **(a)** Five-year Kaplan Meier survival curves for the primary combined end point of death, lung transplant or hospitalization for acute right heart failure of the total PAH cohort (n=197) and **(b)** according to quartiles of hepatic growth factor levels at baseline. **(c)** χ2 of scores and HGF, nerve growth factor (NGF), stem cell growth factor beta (SCGFβ) and stromal cell-derived factor 1 (SDF1) for prediction of the primary end point (death, transplant, or admission for right heart failure) at 3 years in the total PAH cohort (n=197) using Cox regression. Variables were entered in the model using enter mode. The Mayo right heart score was based on the NYHA class, NT-proBNP and RVLS. The Stanford right heart score was based on the NYHA class, NT-proBNP and RVESRI. The REVEAL score was based on the Registry to Evaluate Early And Long-term PAH Disease Management (REVEAL) groups: low, average, moderate high, high, very high. **p<0.01, *p<0.05. NYHA: New York Heart Association; RVESRI: right ventricular end-systolic remodeling index; RVLS: right ventricular longitudinal strain.

Using univariable analysis, HGF and SCGFβ were associated with the primary end point (HR=1.54 [1.24; 1.91], p<0.0001 and 1.26 [1.12; 1.59], p=0.02 respectively), while NGF and SDF1a were not. Kaplan–Meier survival curves for the primary end point according to HGF (p<0.001) and SCGFβ quartiles (p=0.002) are presented in **Figure 3B** and **Figure E2**. HGF added significant incremental value to the Mayo, Stanford and REVEAL scores for prediction of the primary end point at 3 years (p<0.01, **Figure 3C**). Both HGF and SCGFβ were also associated with the secondary end point of death or transplant (HR=1.71 [1.38; 2.12], p<0.0001, and 1.26 [1.01; 1.59], p=0.02 respectively).

### Absence of biomarker of longitudinal cardiac remodeling

Among the total population, 174 patients had a follow-up echocardiogram at 1 year (median time to follow-up study was 1.03 [0.85; 1.35] years). None of the 4 RHMP biomarkers at baseline were found to be associated with changes in right heart remodelling, irrespective of the therapy initiated between studies. Similarly, the Stanford score (p=0.50), Mayo score (p=0.99) and NT-proBNP (p=0.06, OR=1.85 [0.98; 3.49]) at baseline did not associate with changes in right heart remodelling at follow-up.

### Human RV tissue-level expression of RHMP biomarkers

Protein levels of HGF and its receptor c-Met were higher in RV specimens from PAH patients than controls (**Figure 4**). The levels of NGF were decreased and its receptors TRKA increased in PAH as compared to controls.

**Figure 4.**
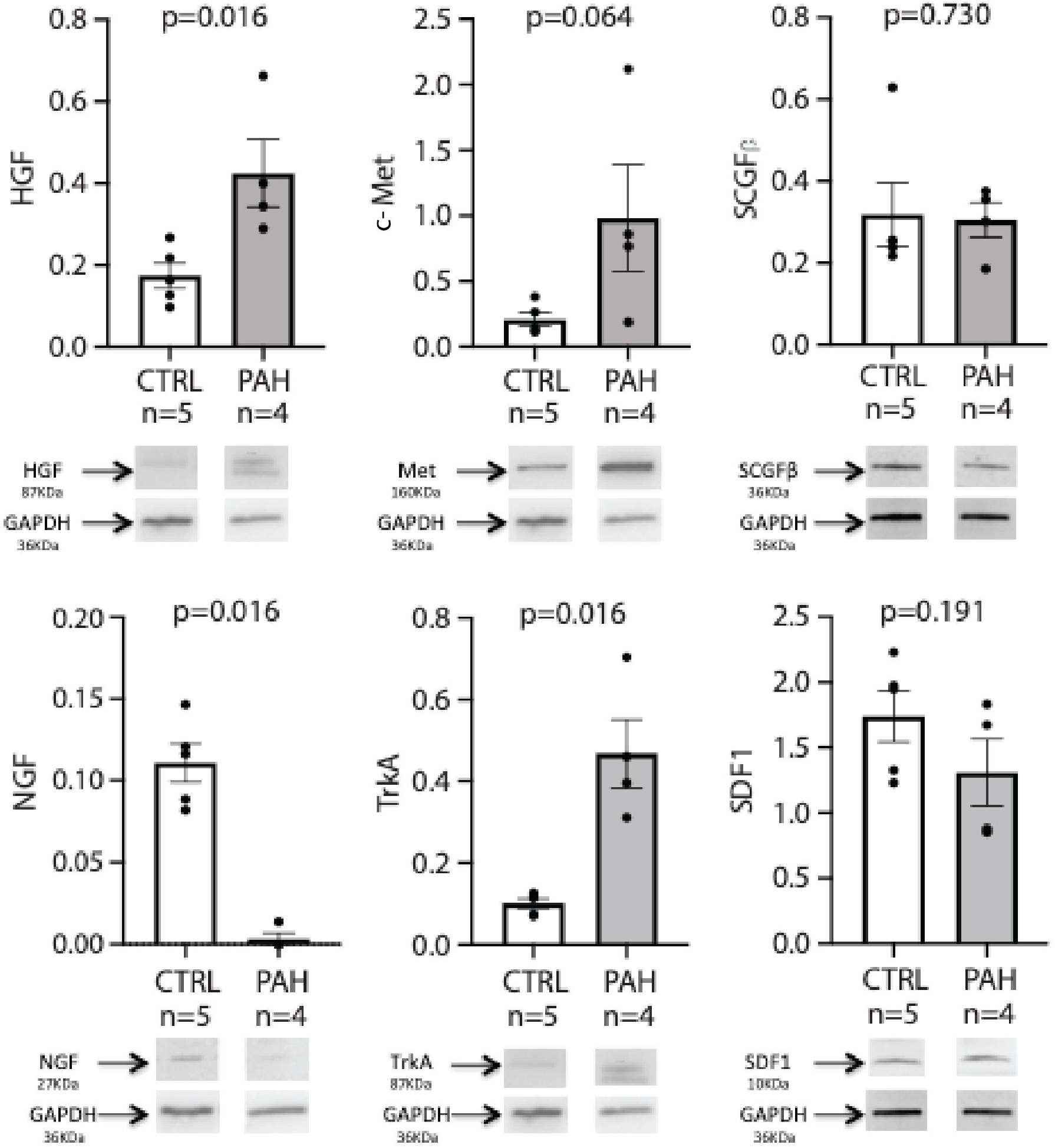
Immunoblot of the protein expression of hepatic growth factor (HGF), its receptor c-Met, Nerve growth factor (NGF) and its receptor TRKA, Stem Cell Growth Factor beta (SCGFb) and SDF1 in the right ventricle of patients with pulmonary arterial hypertension (PAH) and controls, and quantification of the signal. Each bar represents the mean ± SEM signal intensity of Western Blots lysate from right ventricular samples from 4 PAH undergoing heart-lung transplant and 5 control patients (heart donors), which were compared using Student t-test.

In patients with PAH, RV fibrosis level assessed by the % area of Picro-Sirius Red staining was 11.5 [8.8-15.7]%, similar across PAH etiology (**Figure E3** and **Supplementary methods**). **Figure 5** and **Figure E4** show the localization of the 4 biomarkers and receptors in RV samples from the patient with idiopathic PAH. Both PAH patients (idiopathic and BMPR2 mutation) exhibited qualitatively similar protein RV expression. c-Met colocalized with cardiomyocytes, smooth muscle cells, endothelial cells and fibroblasts markers, and to a lesser extent with macrophages. Punctiform expression of HGF was observed in a vesicle pattern, more abundantly in cardiomyocytes, smooth muscle cells, and to a lesser extent in macrophages and fibroblasts. Both TRKA and NGF colocalized with cardiomyocytes, smooth muscle cells and macrophages, while TRKA additionally colocalized with fibroblasts and NGF with endothelial cells (**Figure E4**). SCGFβ was present with cardiomyocytes, smooth muscle cells and fibroblasts, and SDF1 was foudn in cardiomyocytes, smooth muscle cells, endothelial cells and macrophages.

**Figure 5.**
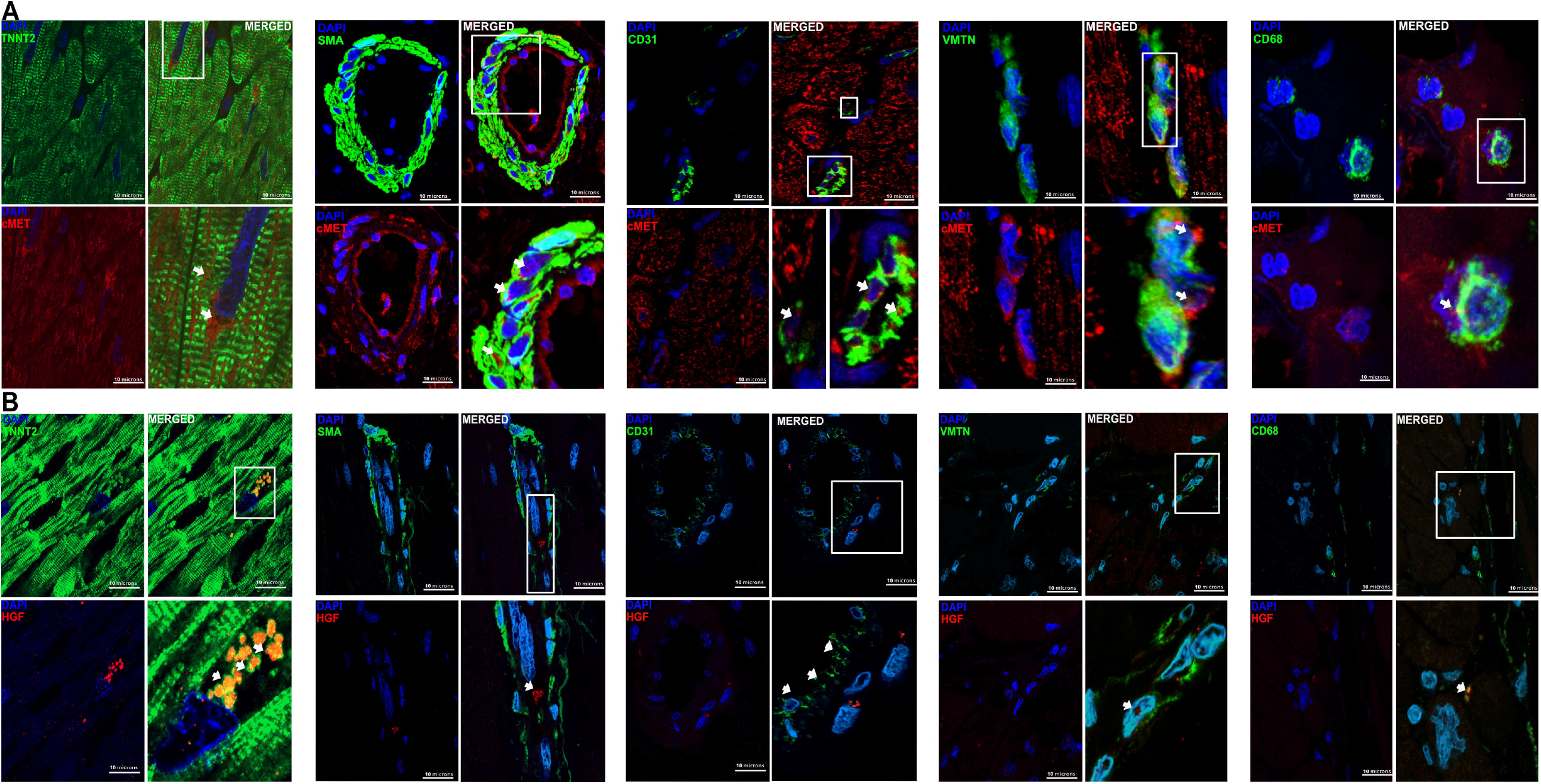
Localization of c-Met (A, red) and its ligand HGF (B, red) in right ventricular biopsies from a patient with idiopathic pulmonary arterial hypertension by immunostaining, double-labeled with either TNNT2 troponin (cardiomyocyte), α-SMA (smooth muscle cell), CD31 (endothelial cells), vimentin VMTN (fibroblast) or CD68 (macrophage) in green, from left to right. No immunoreactivity was seen in cells incubated with the secondary antibody but no primary antibody. Scale barLJ=LJ10 μm. DAPILJ=LJ4′,6′-diamidino-2-phenylindole.

### Altered RV expression of growth factors *Hgf, c-Met, Ngf and Scgf*β in PAB mice

Five weeks after PAB or sham surgery, the RNA expression of *Hgf, c-Met and Ngf* were significantly increased whereas the expression of *Scgf*β was significantly decreased in RV homogenates from PAB compared to shams (**Figure 6**).

**Figure 6.**
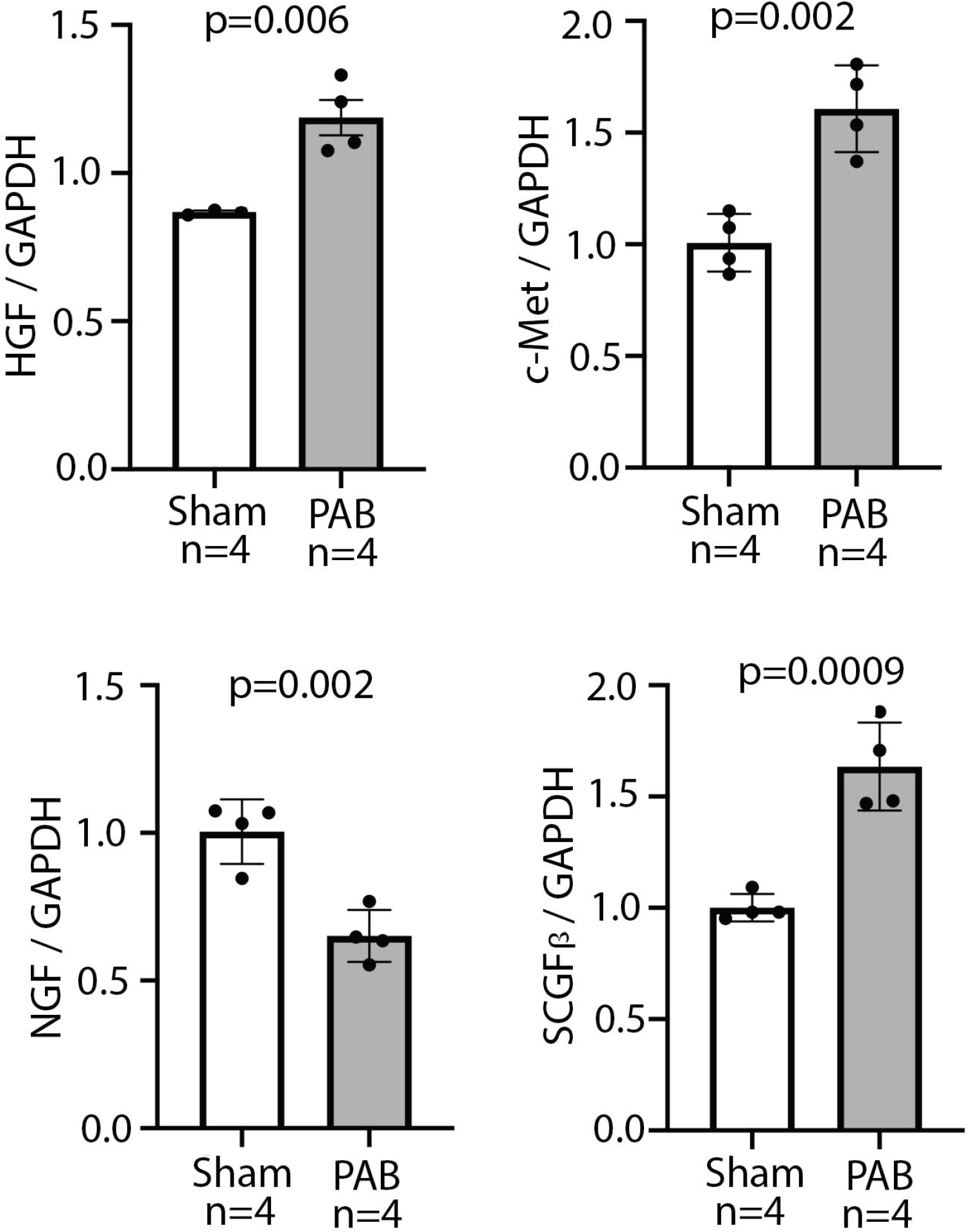
Gene expression of HGF, c-Met, NGF and SCGFb in the RV of pulmonary artery banding mice versus shams. Male C57BL/6 mice (12-14 weeks of age) were anaesthetized and PAB was performed with a non-absorbable silk suture. Five weeks after PAB surgery, right ventricular tissue was collected for gene expression analysis. The RNA expression of *Hgf, c-Met and Ngf* were significantly increased whereas the expression of *Scgf*β was significantly decreased in right ventricular homogenates from PAB compared to Sham control mice. Data are presented as mean±SEM (n=4/group). Student’s 2-tailed, unpaired t-test.

## DISCUSSION

This targeted proteomics study identifies four circulating cytokines associated with RHMP, independently from pulmonary hemodynamic severity. HGF emerges as a strong predictor of clinical worsening at 3 years, incremental to the REVEAL score. The protein expression of c-Met and HGF is increased in the RV of deceased PAH patients in cardiomyocytes, smooth muscle cells and endothelial cells. The PAB mouse model further confirms increased RV gene expression of *Hgf* and *c-Met*, suggesting PAB to be a valid model to further mechanistically study the involvement of this pathway in RV adaptation to PH over time, and answer the question of whether increasing HGF expression represents a beneficial or deleterious response; information potentially useful from a therapeutic aspect. The study originality resides in its design (going from supervised bedside -omics screening to the target organ and bench analysis), which is a novel approach to the pressure-overloaded right heart. This approach has allowed us to use the blood as a window to right heart failure in PAH, leading to the identification of HGF/c-Met as a potential signaling pathway of interest.

HGF is a mesenchymal-derived pleiotropic cytokine binding to its tyrosine kinase receptor c-Met (12). The HGF/c-Met signaling pathway promote angiogenesis, cardiomyocytes survival and inhibit fibrosis experimental myocardial infarction and left cardiomyopathy models (13, 14) and inhibit vascular permeability and inflammation (15)(16). Despite the cardioprotective effects of HGF on the left heart, several studies have paradoxically reported the association between high circulating levels of HGF and left ventricular remodeling, dysfunction or adverse outcomes in several pressure-overloaded cardiac diseases (hypertension and aortic stenosis), or advanced ischemic left heart failure (17–19).

Evidence on the molecular role of HGF/c-Met in the right heart is sparse. High levels of HGF have been reported in small cohorts of patients with scleroderma-associated (20) or idiopathic PAH in whom HGF correlated with MPAP (21). Our study further demonstrates the association of high plasma HGF levels with RHMP beyond pulmonary disease severity. Including two cohorts differing in terms of demographics, prevalence/incidence repartition, and functional severity strengthens the results, suggesting that the biomarkers identified are associated with RV adaptation across PAH etiology. High HGF levels were strongly associated with outcome in our cohort, adding incremental value to the REVEAL risk score, suggesting the timely role of the HGF/c-Met axis on the right heart. None of the 4 RHMP biomarkers but also none of the right heart scores or NT-proBNP levels (validated prognostic markers) at baseline were associated with right heart remodeling improvement at 1 year, irrespective of the therapy initiated, illustrating the distinction between prognostic markers and markers predictive of response to therapy. Future studies should investigate whether changes in biomarkers levels from baseline to 1 year would correlate with changes in right heart remodeling.

While the origin of increased HGF plasma levels in PAH remains to be fully elucidated (increased production of HGF from damaged myocardium, or another organ and/or alterations in HGF systemic clearance), we report the increased protein expression of HGF and c-Met in the RV of patients with PAH with end-stage right heart failure requiring a heart-lung transplant, across PAH etiology. Another original finding is the localization of these markers in the RV of patients with PAH, suggesting the pleiotropic effect of HGF/c-Met as previously demonstrated in the left ventricle (12). The other markers did not show this increased expression in RV samples from patients with PAH as compared to controls, although limited by the number of samples available, which warrants further investigation.

Animal models bring some insights into the potential origin of circulating HGF levels. Increased HGF plasma levels at 2 weeks have been reported in rats with monocrotaline-induced PH, associated with increase in RV mRNA expression of *Hgf* and *c-Met* at 4 weeks, but not in the liver, suggesting a cardiac rather than hepatic origin of plasma HGF (22). Our PAB model further demonstrates the increased gene expression of *Hgf* and *c-Met* in RV homogenates from PAB compared to shams, suggesting that increased circulating levels of HGF in peripheral blood and in the right ventricles of patients with PAH could reflect an increased HGF production in the RV. These data though do not allow the conclusion as to whether increased HGF production in the RV reflects a compensatory attempt of the RV to adapt to an increased afterload or whether it represents rather a harmful response of maladaptation. The overexpression of HGF have also been reported in endothelial cells from PAH patients (23).

The potential beneficial role of HGF as a therapy in PAH has been discussed (24). HGF increases expression of the Bone Morphogenetic Protein Receptor 2, a pathway mutated in the familial form of PAH and downregulated in non-genetic forms (25), and its activation is of benefit PAH and RV failure (26). Several animal models (e.g. rats with monocrotaline-induced PH, rabbits with shunt flow-induced PH) have shown that exogenous HGF or *Hgf* gene transfection reduced the development of PH, which was associated with less marked right heart hypertrophy and lower inflammatory profiles than shams (27–30). However, the timely and direct effect of the HGF/c-Met axis activation in the right heart remains to be elucidated, and here the PAB model might be of particular value to explore the dynamic changes in plasma and tissue HGF expression over time and potential reversibility with de-banding (10).

### Study limitations

The main limitation derives from its single-center design. The fact that the two cohorts were collected and analyzed at two different time points, excluding the risk of batch effect in the proteomics analysis, contributes to increasing the confidence in the results.

In conclusion, high plasma levels of HGF are associated with RHMP in PAH, independent of pulmonary disease severity, and are incremental to the REVEAL score for prediction outcomes. As both RV HGF-cMet protein and gene expression are increased in PAH, this pathway warrants further exploration as a potential RV-specific therapeutic target.

## Data Availability

Due to the PHI sensitivity of clinical database, the dataset is not available online.

## ACKNOWLEDGMENTS

The authors would like to thank the Vera Moulton Wall Center of Pulmonary Hypertension at Stanford and the Stanford Cardiovascular Institute for their funding support. The authors would like to thank Andrew Hsi (Stanford University, CA, USA) for Stanford database management, Catherine Rucker-Martin PhD (from Marie Lannelongue Hospital - Paris Sud Paris Saclay University, France) who highly contributed to the immunofluorescence experiments, Lilia Lamrani, MS (Marie Lannelongue Hospital - Paris Sud Paris Saclay University, France) for her help with clinical data collection and Jean-Baptiste Michel, MD, PhD (Paris VII University, France) for facilitating the procurement of controls cardiac biopsies.

## ABBREVIATIONS LIST

HGF: hepatic growth factor
MPAP: mean pulmonary arterial pressure
NGF: nerve growth factor
NT-proBNP: N-terminal pro brain natriuretic peptide
NYHA: New York Heart Association
PAB: pulmonary arterial banding
PAH: pulmonary arterial hypertension
PAWP: pulmonary arterial wedge pressure
REVEAL: registry to evaluate early and long-termPAH disease management
RHF: right heart failure
RV: right ventricle
RVESAI: RV end-systolic area index
RVESRI: RV end-systolic remodeling index
RVFAC: RV fractional area change
RVLS: right ventricular longitudinal strain
TAPSE: tricuspid annular plane systolic excursion
SDF1: stromal derived factor-1
SCGFβ: stem cell growth factor beta

